# Optimizing stimulation parameters for anterior thalamic nuclei deep brain stimulation in epilepsy: A randomized cross-over trial

**DOI:** 10.1101/2025.01.22.25320034

**Authors:** Juan Luis Alcala-Zermeno, Gamaleldin Osman, Jayawant N. Mandrekar, Keith Starnes, Nicholas M. Gregg, Gregory Worrell, Brian N. Lundstrom

## Abstract

**Objective:** The effects of brain stimulation for diseases like epilepsy are delayed, making stimulation optimization difficult. The parameters for anterior thalamic nuclei (ANT) deep brain stimulation (DBS) for focal drug-resistant epilepsy (DRE) management are often restricted to those used in the SANTE landmark trial. There is little evidence regarding effective alternatives, and low frequency stimulation is typically neglected. We prospectively compare a widely differing stimulation parameter set to typical settings.

**Methods:** This randomized, modified cross-over, open trial compares the efficacy and safety of an alternative set of parameters using continuous low frequency stimulation with longer pulse-width (cLFS), (7 Hz, 200 msec, continuous) compared to SANTE’s intermittent high frequency stimulation with a short pulse width (iHFS), (145 Hz, 90 msec, cycling 1 min on/5 min off). After 3 months on a randomly assigned first set, patients are switched to the other settings, unless seizure free. Patients are re-evaluated after 3 more months at which point they can either remain on the same settings or switch back.

**Results:** Sixteen patients with a median baseline seizure frequency of 13.8 sz/month (IQR 2.7-22.8) were included in the analysis. At last-follow up, ANT-DBS significantly reduced median seizure frequency (45%, IQR 3 - 80%; p = .04). Both iHFS (33%, IQR 0 - 65; p = .02) and cLFS (72%, IQR 30 - 79; p = .001) significantly reduced median seizure frequency. cLFS showed improved median seizure frequency reduction compared to iHFS (p = .03) and was not associated with any moderate or severe adverse effects.

**Significance:** Results support cLFS for ANT-DBS as a safe and effective alternative to typical iHFS parameters. Broadly, stimulation with widely differing parameters sets may be as effective or even more effective than typical stimulation parameters.

## Introduction

Brain stimulation is increasingly used to treat a wide array of neurological diseases. For some conditions, like epilepsy, the effect of stimulation is not immediate, making stimulation optimization difficult and the extent to which optimization can improve outcomes is largely unknown. Deep brain stimulation (DBS) in an invasive neuromodulation strategy available to treat drug resistant epilepsy (DRE) that is not amenable to resective epilepsy surgery.^1^ Current implantable pulse generators (IPGs) can provide DBS therapy in a multitude of combinations of amplitude, frequency, pulse width, cycling, and electrode configuration. Yet, the stimulation parameters used for anterior thalamic nuclei (ANT) DBS in focal DRE are often restricted to a narrow set of parameters closely related to the SANTE landmark trial settings, which consist of single cathodal referential stimulation using 145 Hz frequency, 90 msec pulse width, and cycling stimulation 1 min on and 5 min off.^2^ Unfortunately, not all patients respond to the SANTE parameters and the evidence for invasive ANT stimulation in epilepsy using widely differing alternative settings is sparse, and mainly restricted to animal models^3^ or small retrospective case series reporting the outcomes of corticothalamic responsive neurostimulation (RNS)^4^ or fornix DBS,^5^ which is a critical ANT afferent through direct fornicothalamic and indirect mammillothalamic projections.^6^ A neglected strategy in epilepsy DBS that is sometimes described as worsening seizures is low frequency stimulation, which can reduce seizure frequency when targeting cortex using RNS^7^ or chronic subthreshold stimulation (CSS).^8^ We want to prospectively compare stimulation that notably differs in frequency, pulse width, and duty cycle (continuous vs. intermittent) from standard parameters. Specifically, our primary objective is to prospectively compare an alternative set of DBS parameters using **c**ontinuous **L**ow **F**requency **S**timulation (cLFS) with the SANTE trial parameters which use **i**ntermittent **H**igh **F**requency **S**timulation (iHFS) with regard to seizure frequency reduction (SFR). Secondarily, we aim to characterize the effects of cLFS and iHFS on patient-perceived outcomes.

## Materials & Methods

This prospective, single center, randomized, cross-over trial (NCT06617845) was approved by the Mayo Clinic Institutional Review Board to study two sets of stimulation parameters in patients implanted with bilateral ANT-DBS between 2021 and 2023 for management of DRE (Figure *1*). Chronic stimulation leads were implanted using a posterior transcortical approach with direct ANT targeting as described previously by our group.^9^ After implantation, patients were randomized to either iHFS (145 Hz frequency, 90 msec pulse width, and cycling 1 min on and 5 min off) or cLFS (7 Hz frequency, 200 msec pulse width, and continuous stimulation). The choice of low frequency stimulation using 7 Hz was relatively arbitrary, although our group had prior experience with this frequency and theta oscillations are prevalent within the limbic system.^7, 10^ Low frequency stimulation was delivered continuously and with a longer pulse width, which reduces the difference in total charge delivered per day as compared to typical SANTE parameters. Both iHFS and cLFS were programmed using referential stimulation with two cathodes (to minimize any targeting differences). The choice of cathodes was guided by post-implant imaging. In general, we used the two most distal electrodes per lead as long as pre and postoperative image co-registration demonstrated placement within ANT (Supplementary material). For each parameter set, stimulation current was started at 2-4 mA per lead, as tolerated. Participants were instructed to go up by 1-2 mA after a month to a target current of 5-6 mA per lead, as tolerated. To assess if the amount of stimulation was different between parameters, we calculated the charge delivered per day in Coulombs (C) using the following formula:

**Figure 1.**
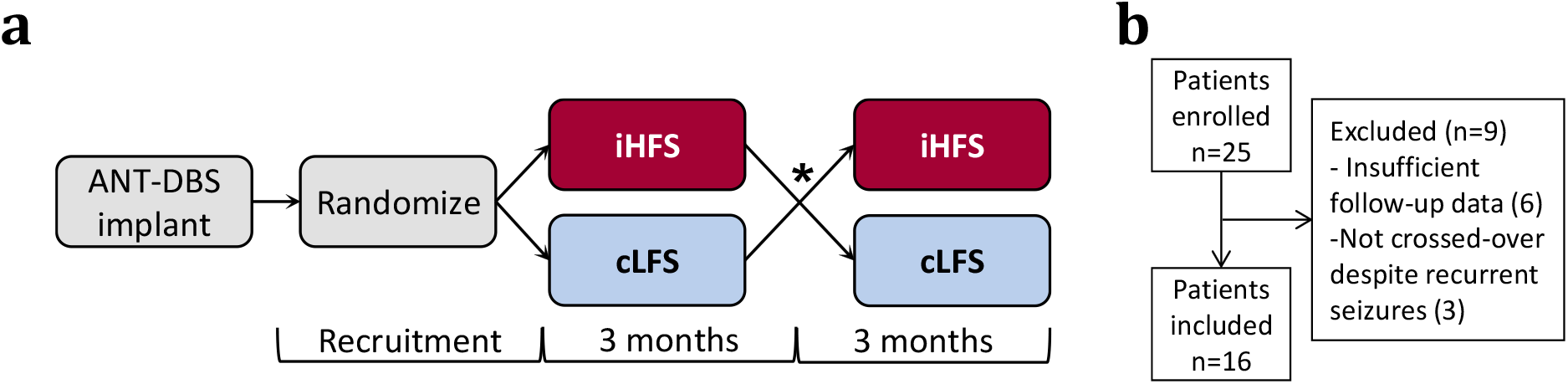
**a)**: Schematic of trial timeline. Two weeks from implantation, patients were randomized to either iHFS or cLFS for 3 months. On the irst visit, patients were crossed over to cLFS or iHFS for 3 months. * If patients were seizure-free on the irst stimulation settings they were not crossed over. **b)**: Patient participation

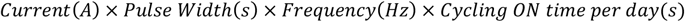

At least two weeks were allowed between implantation and randomization to account for any implant effect. Baseline seizure frequency was determined as seizures per month in the three months preceding each assessment. The protocol included two mandatory seizure frequency assessments. The first seizure frequency was evaluated three months after starting stimulation. Patients were then crossed over to the other parameter set (unless they were seizure-free). A second seizure frequency assessment was performed three months later. Patients were then managed per physician’s preference with an optional third seizure frequency assessment at least three months later.

Patient perceived outcomes were evaluated at baseline and at every assessment visit using 1-10 numeric analog scales to identify seizure severity, life satisfaction, and quality of sleep. Implantation-related adverse events (AE) and stimulation-related side effects (SE) were assessed at baseline and at every visit (Supplementary forms 1 and 2). Antiseizure medication (ASM) management followed primary neurologist/epileptologist’s preference. Continuous variables are reported as median with interquartile range (IQR). Non-parametric testing was used for comparisons (Mann-Whitney U test or Wilcoxon matched-pairs signed rank test, as appropriate). Significance was established at p < .05. All analyses were performed in GraphPad Prism version 10.2.3 for Windows, (GraphPad Software, Boston, Massachusetts, USA). DBS-lead visualization and localization was performed through *Lead-DBS* package v.2.5.3^11^ and Lead-Group addition^12^ using the Krauth/Morel thalamic atlas adjusted for use in Montreal Neurological Institute (MNI) 2009b asymmetric template space (*Figure 2*).^13^

**Figure 2.**
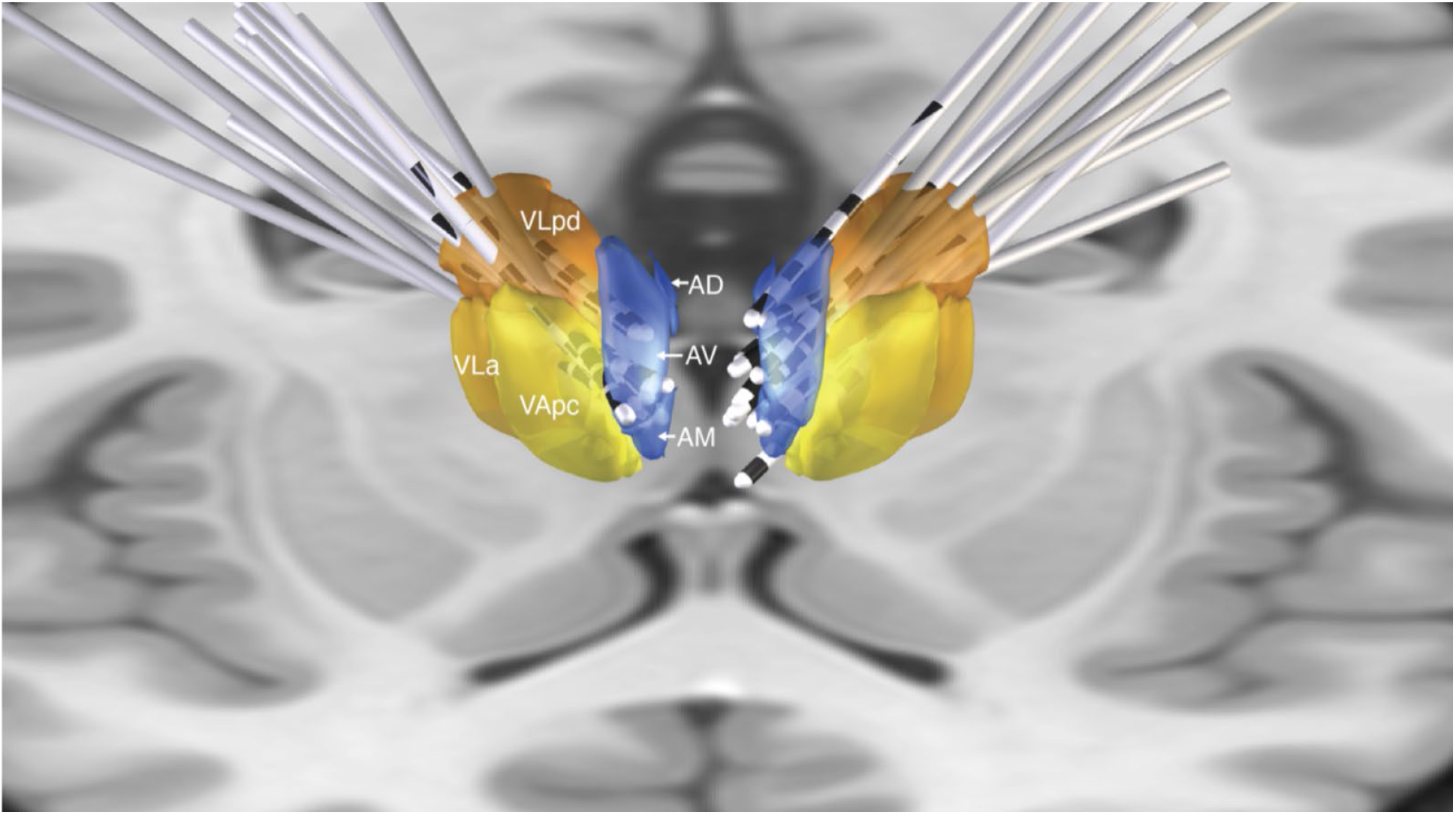
Group lead reconstruction of all DBS implants with respect to their thalamic targets. AD – anterodorsal thalamic nucleus, AM - anteromedial thalamic nucleus, AV - anteroventral thalamic nucleus, VApc - ventral anterior thalamic nucleus pars parvocelullaris, VLa – ventral lateral thalamic nucleus pars anterior. VLpd – ventral lateral thalamic nucleus posterior dorsal.

## Results

Sixteen patients completed the trial with 13 completing an optional third follow-up visit (Figure *1*). Baseline characteristics of the participants are delineated in Table 1. Median baseline seizure frequency was 13.8 seizures per month (sz/month) (IQR 2.7 - 22.8), trial follow-up time (two assessments) was 30 weeks (IQR 26 - 35), and total follow-up time (three assessments) was 49 weeks (IQR 35 - 56). At the first follow-up visit, patients 2 and 10 became seizure-free on iHFS and cLFS, respectively, so they did not cross over and remained on the same parameters for the second assessment. For the patients that crossed over, there was no difference in time spent on iHFS vs cLFS (median 14 wk, IQR 13 - 21 vs 14 wk, IQR 11 - 15; W = -32, p = .3). At last follow up (median 49 weeks, IQR 35 - 56), ANT-DBS significantly reduced median seizure frequency (48%, IQR 3 - 84, W = 79; p = .04) (Figure *3*). Of the 13 who completed the optional third assessment, four of them (31%) remained on iHFS, and 9 (69%) on cLFS (Supplementary figure 1).

**Figure 3.**
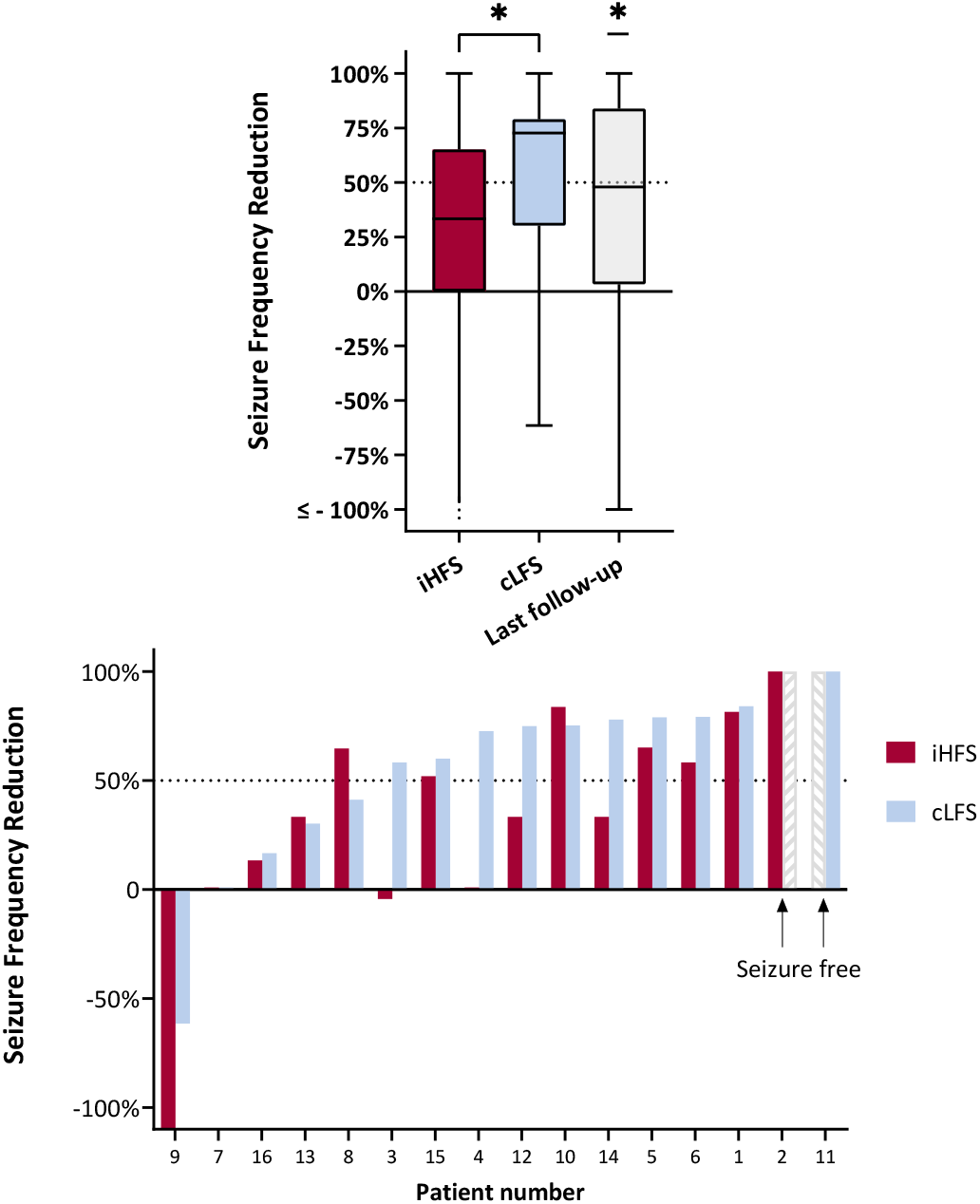
**Upper panel** - Median SFR according to stimulation parameters at last follow up. iHFS, cLFS and ANT-DBS at last follow up all demonstrated a signiicant SFR. When comparing iHFS vs cLFS (Wilcoxon signed-rank test), cLFS showed a signiicantly improved SFR than iHFS. **Lower panel** - Individual seizure frequency reduction with iHFS and cLFS from worst (lower left) to best (upper right). cLFS, continuous low frequency stimulation, iHFS, intermittent high frequency stimulation

**Table 1.**
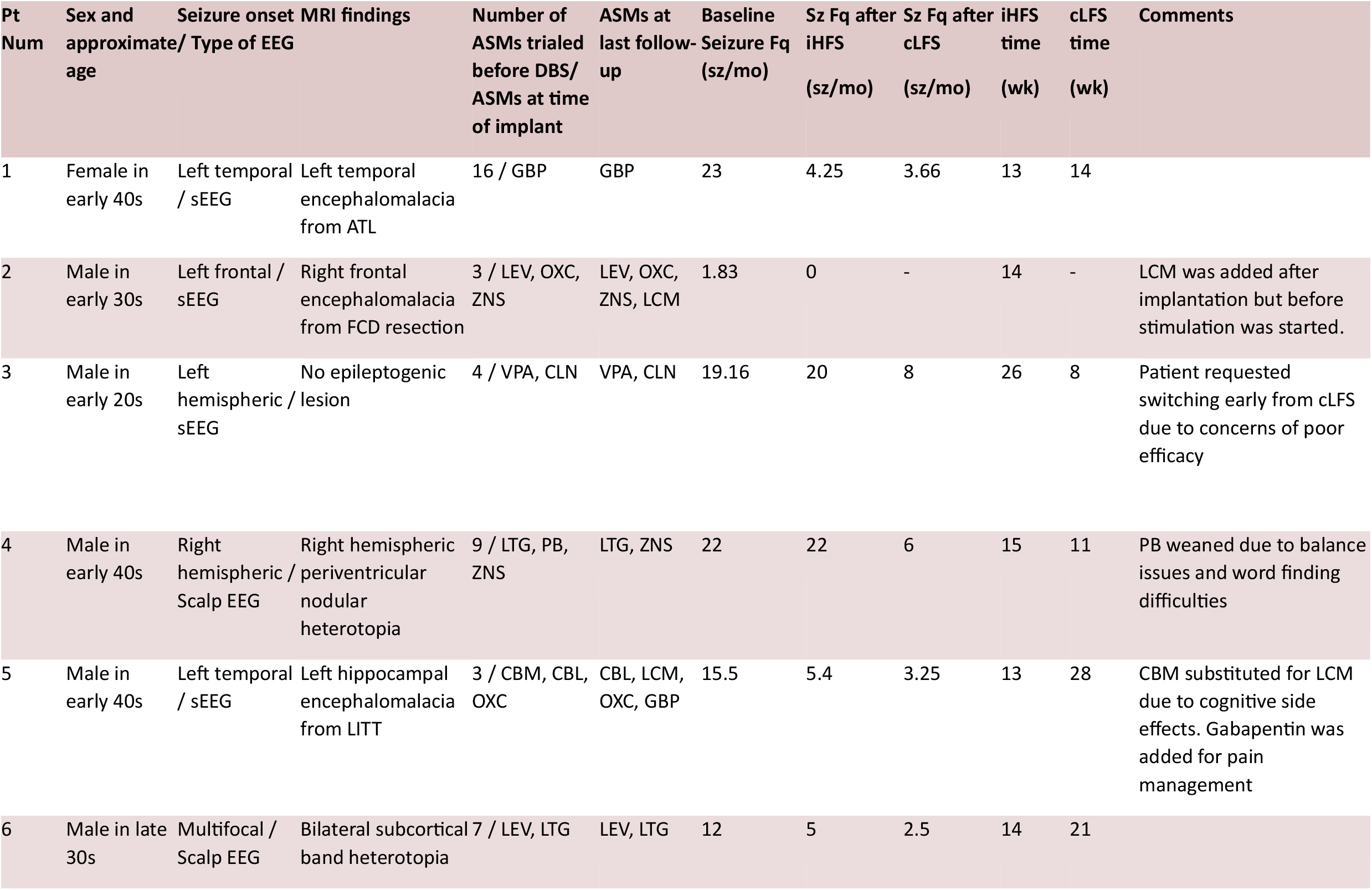
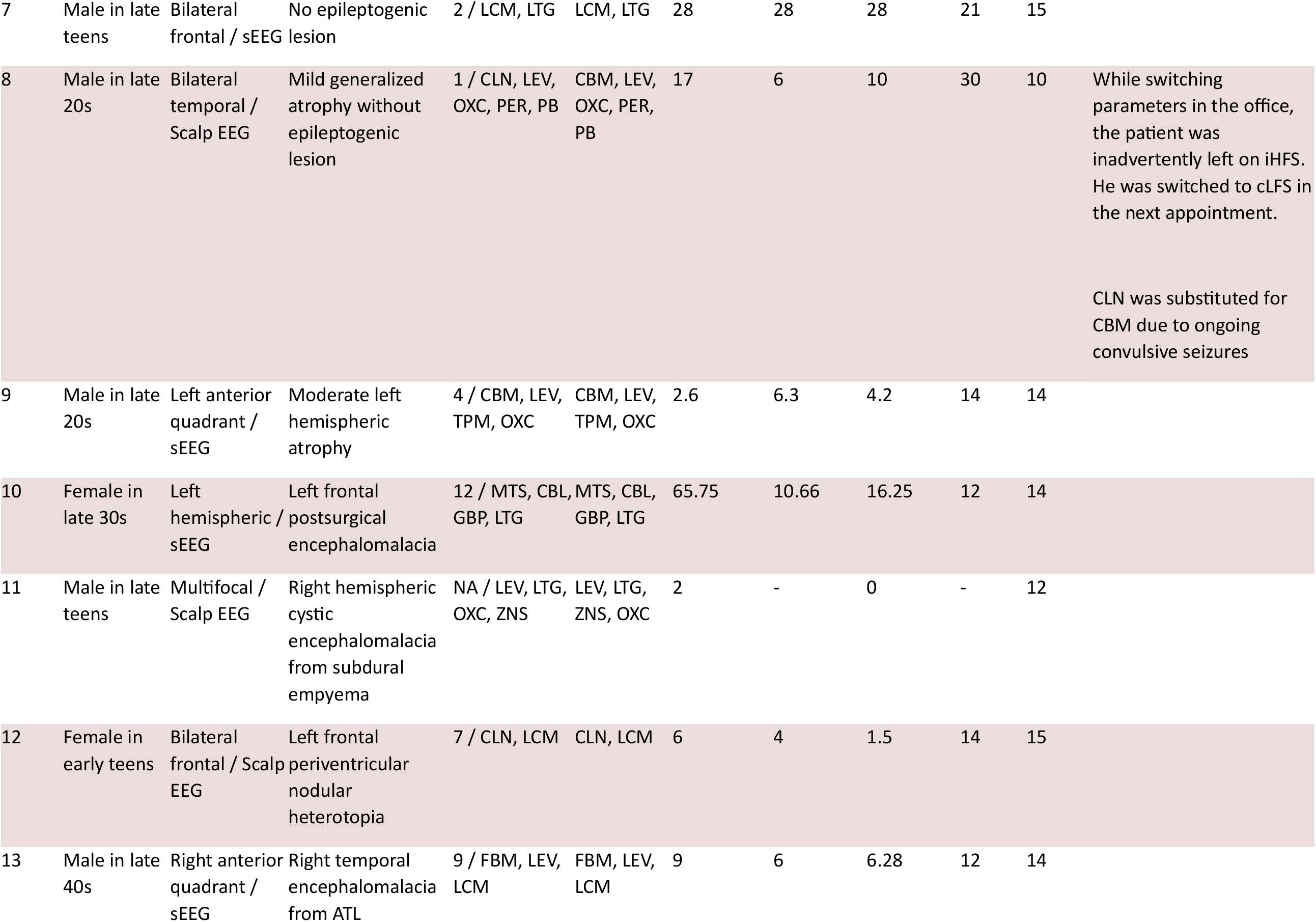
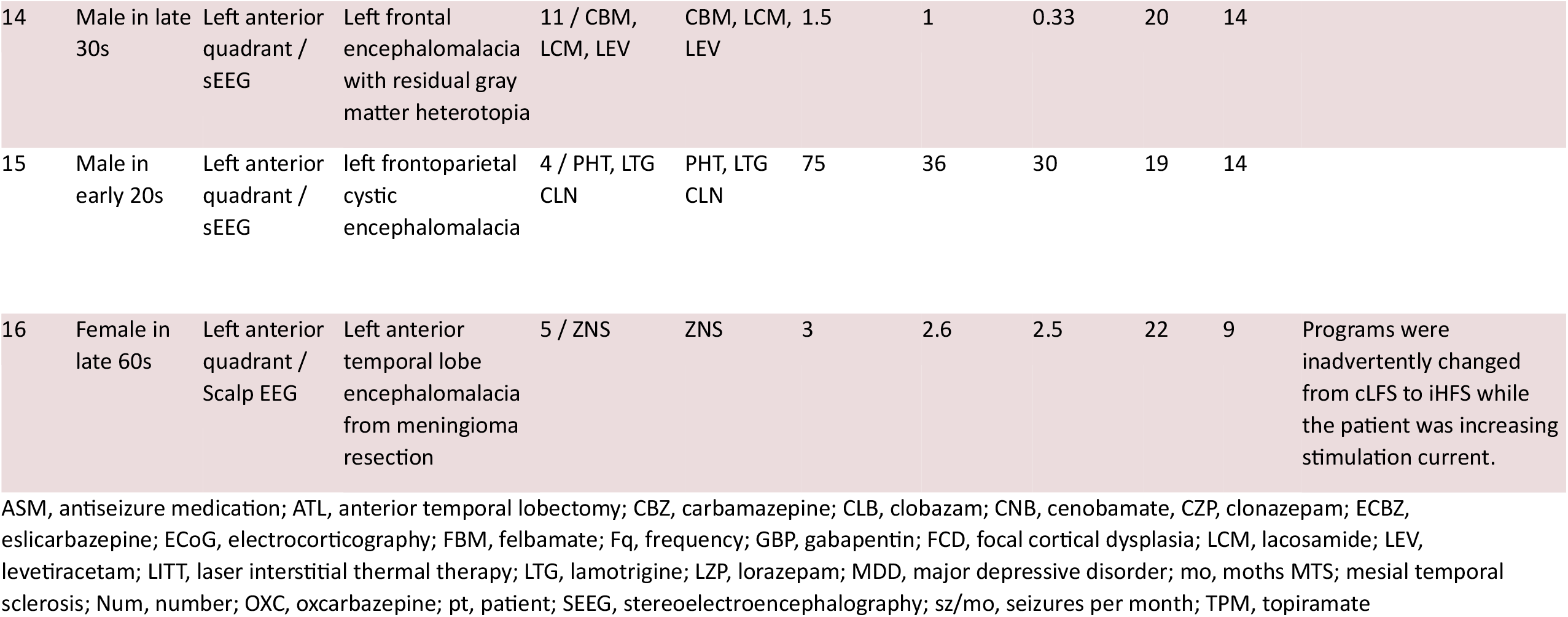
Baseline characteristics.

### Primary outcome

Both iHFS (33%, IQR 0 - 65, W = 81; p = .02) and cLFS (72%, IQR 30 - 79, W = 105; p = .001) reduced median seizure frequency. cLFS had improved median SFR compared to iHFS (W = 61; p = .03).

### Secondary outcomes

At last follow-up, ANT-DBS did not improve seizure severity (0%, IQR -11 to 25, W = 12; p = .8), life satisfaction (6%, IQR 0 - 39, W = 60; p = .08), or quality of sleep (0%, IQR 0 - 22, W= 35; p = .3) (Supplemental figure 2). Ten patients (62%, IQR 39 - 82) reported AEs related to hardware implantation, with one patient having an AE of moderate severity (requiring dedicated medical evaluation and/or non-invasive intervention) due to a small intraparenchymal bleed around a DBS lead associated with mild facial asymmetry and upper extremity drift that resolved spontaneously after 24 hours. Eleven patients (69 %, IQR 45 - 86) reported stimulation-related SEs, with only two patients having moderate severity ones, both of which were mood side effects while on iHFS that required parameter adjustment (Supplementary material and supplementary Figure 4). There were no statistical differences in the proportion of patients who showed at least a 50% SFR (responders) to iHFS (44%) or cLFS (63%) (X^2^ = 3.7, df = 4, p = .5) or at least a 75% SFR (super-responders) to iHFS (18%) or cLFS (44%) (X^2^ = 8.4, df = 4, p = .08).

Charge delivered per day was approximately 36% lower in the cLFS group compared to the iHFS (1.88 C/d, IQR 1.22 - 1.88 vs 1.21 C/d, IQR 0.79 - 1.452; p = .003).

## Discussion

The results of this randomized cross-over open-label clinical trial of 16 patients suggest that ANT-DBS can be effective in reducing seizure frequency not only with the typical iHFS parameters used in the SANTE trial but also with an alternative stimulation paradigm using continuous low frequency stimulation with longer pulse widths with a similar safety profile.

The overall median 48% SFR observed at last follow up is consistent with SANTE trial’s unblinded phase where 41% and 56% SFRs were observed at 13 and 25 months, respectively.^2^ When looking at iHFS, we observed a less impressive median SFR of 33% (with 3 months iHFS stimulation) compared to the 40.4% observed in the SANTE trial at 3-4 months.^2^ Although this may represent random variability, it could also be explained by a higher prevalence of severe cases, as patients in this study had a higher number of ASMs at baseline (62% of patients taking ≥3 ASM in this trial vs. 40% in the SANTE trial), and most had diffuse or multifocal onsets (63% vs 9% in the SANTE trial). This has been seen in other real-world DBS-ANT studies.^14, 15^

Anterior thalamic nuclei stimulation produces different activation patterns in the default mode network and limbic structures depending on its frequency.^16^ Low frequency stimulation may be associated with decreased activation of the limbic network, which may better desynchronize ictal rhythms and improve seizure frequency reduction.^17^ However, there is individual variability as some patients responded better to low frequency parameters and others to high frequency, a phenomenon that has been demonstrated in cortical and corticothalamic RNS where both low and high frequency stimulation have been effective.^4, 7^ The lower stimulation frequency may not be the only difference accounting for the efficacy of our alternative parameter set as it had a longer pulse width and was delivered continuously (no cycling). Interestingly, the overall charge delivered per day was lower with cLFS compared to the SANTE parameters, suggesting that effectiveness is not tightly related to charge delivery. The individual contributions of pulse width, cycling, and stimulation frequency apart from charge delivery remains to be elucidated.

Our study adds to the increasing evidence supporting the safety of low frequency thalamic stimulation for at least some parameter settings.^4, 18-20^ Specifically, we have demonstrated the safety of 7 Hz stimulation in ANT. However, the generalizability of our findings is somewhat limited as we did not attempt lower frequencies to exclude the triggering of seizures seen with 3 Hz centromedian thalamic nucleus stimulation of DBS-naïve patients with absence seizures using high voltage in a single case series.^21^

On AEs related to hardware implantation and stimulation-related SE, we did observe a clinically minor hemorrhage (6%), which is similar to the 4.5% observed in SANTE.^2^ We also noted a higher incidence of mood side effects (31% vs. 14.8% in SANTE), however all of the mood changes were self-limited and only patients who received iHFS required stimulation parameter modifications. This may be related to the different limbic activation patterns seen with high frequency stimulation vs. low frequency.^22^ Limitations include the start of stimulation two weeks after implant, which is shorter than the four weeks used in the SANTE trial. However, any degree of implant effect would affect both stimulation settings equally and not affect any comparisons. Another limitation is the lack of washout period between the two stimulation sets. Given the effectiveness of low frequency stimulation in other contexts,^20, 23^ we were concerned about stopping an effective therapy. In addition, since patients started with a randomly chosen parameter set (the order of parameter sets were evenly split within our patient cohort), any carryover effects would lessen rather than increase any observed effects size. The only exception to this would perhaps be with a priming effect, where the order of stimulation sets itself (i.e. cLFS and then iHFS compared to iHFS and then cLFS) represent the comparison of interest. However, although limited statistically by small numbers, we see no evidence of a priming effect as there was no median SFR difference in patients who started on one set vs the other. Finally, three months on each parameter set is limited and a longer time may have been ideal. However, three months were used in the landmark trials and practically these parameter sets are only two of an infinite number, and a longer period would delay further optimization.

## Conclusion

This trial suggests that ANT-DBS using cLFS can be an effective and safe alternative to typical high frequency parameters for management of DRE. More importantly, results suggest that stimulation optimization can be effective for individual patients and may include a wider variety of stimulation parameter choices than typically considered.

## Acknowledgements

We thank Karla Crockett, Isabelle Meunier, Amy Headlee, and Julianna Ethridge for providing impeccable support as research coordinators.

## Disclosure of conflicts of interest and ethical publication statement

G.W. and B.N.L. are named inventors for intellectual property licensed to Cadence Neuroscience, which is co-owned by Mayo Clinic. B.N.L. waived contractual rights to royalties. G.W., N.M.G., and B.N.L. are investigators for the Medtronic EPAS trial and Medtronic-supported NIH grants (UH3-NS95495 and UH3-NS112826). B.N.L. is an investigator for the Neuropace RNS System Responsive Stimulation for Adolescents with Epilepsy (RESPONSE) study and Neuroelectrics tDCS for Patients with Epilepsy study. Mayo Clinic has received consulting fees on behalf of B.N.L. from Epiminder, Medtronic, Neuropace, and Philips Neuro. Mayo Clinic has received research support and consulting fees on behalf of G.W. from UNEEG, NeuroOne, and Medtronic. G.W. has licensed intellectual property developed at Mayo Clinic to NeuroOne. Neither of the other authors has any conflict of interest to disclose.

## Statements

### Data availability

The data that support the findings of this study are available from the senior author, (B.N.L.), upon reasonable request.

### Funding

No specific funding was received for this study.

### Ethics approval

The Institutional Review Board approved this study under the IRB 20-009654

### Patient consent

Written informed consent was obtained from all patients for the publication of this study.

## Notes

### Competing Interest Statement

G.W. and B.N.L. are named inventors for intellectual property licensed to Cadence Neuroscience, which is co-owned by Mayo Clinic. B.N.L. waived contractual rights to royalties. G.W., N.M.G., and B.N.L. are investigators for the Medtronic EPAS trial and Medtronic-supported NIH grants (UH3-NS95495 and UH3-NS112826). B.N.L. is an investigator for the Neuropace RNS System Responsive Stimulation for Adolescents with Epilepsy (RESPONSE) study and Neuroelectrics tDCS for Patients with Epilepsy study. Mayo Clinic has received consulting fees on behalf of B.N.L. from Epiminder, Medtronic, Neuropace, and Philips Neuro. Mayo Clinic has received research support and consulting fees on behalf of G.W. from UNEEG, NeuroOne, and Medtronic. G.W. has licensed intellectual property developed at Mayo Clinic to NeuroOne. Neither of the other authors has any conflict of interest to disclose.

### Clinical Trial

NCT06617845

### Author Declarations

This prospective, single center, randomized, cross-over trial (NCT06617845) was approved by the Mayo Clinic Institutional Review Board

