## Supplementary material for "Optimizing stimulation parameters for anterior thalamic nuclei deep brain stimulation in epilepsy: A randomized cross-over trial"

### Calculation of lead electrode to anatomical target

DBS-lead localization determined using *Lead-DBS* package v.2.5.3[1] with anatomical referencing as per the Krauth/Morel thalamic atlas adjusted for use in Montreal Neurological Institute (MNI) 2009b asymmetric template space.[2] Regarding proximity, we determined the closest distance to AM and AV as the most relevant target since both are components of the anterior thalamic nuclei (ANT). Please bear in mind that the software used measures distance from contact center to closest vertex of rendered structure, so numbers will be >0 even when a contact is within a structure. Distances <1.5mm in *Lead-DBS* are considered appropriate.

Supplementary table 1 – Individual electrode distance from target

| Patient | Contact | AD | AM | AV | CL | MDpc | VApc | VLa | VLpd | VLpv | MTT |
|---|---|---|---|---|---|---|---|---|---|---|---|
| **1** | 0 | 5.11 | 0.67 | 1.31 | 2.21 | 2.85 | 3.97 | 9.15 | 5.45 | 4.9 | 4.1 |
| | 1 | 3.31 | 1.46 | 0.27 | 0.3 | 1.61 | 2.05 | 6.12 | 2.12 | 1.93 | 1.43 |
| | 2 | 3.9 | 4.75 | 1.04 | 0.28 | 0.94 | 2.58 | 3.74 | 0.63 | 0.37 | 1.99 |
| | 3 | 6.06 | 8.13 | 2.95 | 0.49 | 1.11 | 3.87 | 3.4 | 0.13 | 0.62 | 5.21 |
| | 8 | 6.35 | 1.04 | 0.29 | 1.8 | 3.25 | 0.95 | 6.48 | 2.9 | 2.26 | 1.57 |
| | 9 | 6.11 | 3.95 | 1.87 | 1.72 | 3.23 | 0.55 | 3.19 | 0.19 | 0.19 | 0.81 |
| | 10 | 7.47 | 7.18 | 4.14 | 2.37 | 3.19 | 0.74 | 0.74 | 1.9 | 0.25 | 3.61 |
| | 11 | 9.75 | 10.5 | 6.52 | 2.64 | 3.38 | 3.19 | 0.67 | 1.28 | 0.24 | 6.87 |
| **2** | 0 | 4.37 | 0.57 | 0.57 | 2.08 | 2.84 | 4.2 | 8.77 | 4.89 | 4.49 | 4.02 |
| | 1 | 3.05 | 0.49 | 0.34 | 1.5 | 2.53 | 2.89 | 7.15 | 3.06 | 2.91 | 2.55 |
| | 2 | 2.77 | 2.55 | 0.11 | 1.34 | 2.46 | 1.92 | 5.68 | 1.84 | 2.14 | 2.28 |
| | 3 | 3.49 | 4.62 | 0.22 | 0.87 | 2.27 | 2.34 | 4.64 | 0.88 | 1.22 | 3.32 |
| | 8 | 5.95 | 2.9 | 1.25 | 1.83 | 3.64 | 0.72 | 4.29 | 0.84 | 0.43 | 0.29 |
| | 9 | 6.5 | 5.06 | 2.48 | 2.17 | 3.27 | 0.35 | 2.06 | 1.2 | 0.33 | 1.87 |
| | 10 | 7.73 | 7.31 | 4.2 | 2.82 | 3.64 | 0.38 | 0.38 | 1.75 | 0.19 | 3.81 |
| | 11 | 9.38 | 9.58 | 6.05 | 3.06 | 3.98 | 1.96 | 0.37 | 0.48 | 0.15 | 6 |

| | | | | | | | | | | | |
|---|---|---|---|---|---|---|---|---|---|---|---|
| **3** | 0 | 5.19 | 0.79 | 1.67 | 2.18 | 2.63 | 4.05 | 9.37 | 5.63 | 5.12 | 4.23 |
| | 1 | 3.68 | 0.24 | 0.24 | 0.75 | 1.88 | 3.08 | 7.27 | 3.4 | 2.97 | 2.47 |
| | 2 | 3.2 | 2.28 | 0.29 | 0.64 | 2.13 | 1.3 | 5.34 | 1.28 | 1.24 | 1.06 |
| | 3 | 3.93 | 4.49 | 0.48 | 0.37 | 1.97 | 1.75 | 3.67 | 0.37 | 0.82 | 2.16 |
| | 8 | 3.88 | 0.97 | 0.19 | 1.26 | 2.56 | 2.34 | 6.58 | 2.72 | 2.32 | 1.89 |
| | 9 | 3.96 | 3.06 | 0.24 | 0.24 | 2.32 | 0.69 | 4.47 | 0.63 | 0.39 | 0.39 |
| | 10 | 5.06 | 5.21 | 1.69 | 0.89 | 1.8 | 1.95 | 2.64 | 0.48 | 0.25 | 1.83 |
| | 11 | 6.8 | 7.46 | 3.84 | 1.38 | 2.08 | 2.03 | 1.85 | 1.16 | 0.27 | 3.89 |
| **4** | 0 | 1.01 | 1.84 | 0.72 | 0.72 | 1.22 | 4.36 | 8.16 | 4.17 | 4.18 | 3.96 |
| | 1 | 1.42 | 4.48 | 0.22 | 0.22 | 0.28 | 3.65 | 5.81 | 2.07 | 2.15 | 3.66 |
| | 2 | 3.01 | 7.78 | 0.87 | 0.26 | 0.16 | 4.91 | 5.33 | 0.83 | 2.01 | 5.77 |
| | 3 | 5.49 | 11.3 | 2.56 | 0.27 | 0.76 | 6.63 | 5.95 | 0.27 | 2.71 | 8.97 |
| | 8 | 6.5 | 1.61 | 0.13 | 2.95 | 4.44 | 0.87 | 6.37 | 3.14 | 2.84 | 1.76 |
| | 9 | 5.58 | 4.19 | 0.87 | 1.56 | 3.78 | 0.15 | 3.09 | 0.43 | 0.28 | 0.28 |
| | 10 | 6.81 | 7.02 | 2.94 | 2.52 | 3.47 | 0.18 | 0.18 | 0.24 | 0.18 | 3.51 |
| | 11 | 8.74 | 10.34 | 5.25 | 3.67 | 4.46 | 1.87 | 0.71 | 0.24 | 1.32 | 6.88 |
| **5** | 0 | 1.12 | 4.61 | 0.22 | 2.56 | 3.56 | 5.08 | 8.04 | 4.3 | 4.62 | 5.55 |
| | 1 | 1.07 | 7.21 | 0.17 | 1.87 | 2.73 | 4.47 | 6.7 | 1.96 | 3.54 | 6.45 |
| | 2 | 3.07 | 10.63 | 0.28 | 1.48 | 2.65 | 6.76 | 6.97 | 0.34 | 5.48 | 9.15 |
| | 3 | 6.17 | 14.43 | 2.58 | 1.65 | 4.47 | 8.68 | 8.8 | 0.4 | 6.24 | 12.73 |
| | 8 | 2.86 | 4.11 | 0.23 | 1.23 | 2.17 | 2.81 | 5.41 | 1.71 | 1.96 | 3.47 |
| | 9 | 3.39 | 7.16 | 0.38 | 0.17 | 1.01 | 3.29 | 3.92 | 0.17 | 1.28 | 4.95 |
| | 10 | 4.97 | 10.47 | 2.12 | 0.34 | 1.35 | 4.56 | 4.09 | 0.24 | 2.35 | 7.88 |
| | 11 | 7.51 | 13.96 | 4.55 | 1.88 | 3.05 | 6.75 | 5.48 | 0.14 | 2.2 | 11.31 |
| **6** | 0 | 2.48 | 1.48 | 0.3 | 1.62 | 2.7 | 2.81 | 6.92 | 2.87 | 2.96 | 2.77 |
| | 1 | 2.85 | 4.26 | 0.37 | 0.3 | 0.83 | 1.89 | 3.91 | 0.47 | 0.78 | 1.86 |
| | 2 | 5.48 | 7.5 | 2.26 | 0.28 | 0.54 | 3.12 | 2.71 | 0.3 | 0.31 | 4.44 |
| | 3 | 8.12 | 11.04 | 5.46 | 0.89 | 1.78 | 5.19 | 3.14 | 0.41 | 0.26 | 7.83 |
| | 8 | 5.16 | 1.35 | 0.25 | 2.1 | 3.52 | 1.99 | 6.58 | 3.23 | 2.58 | 1.84 |
| | 9 | 4.02 | 3.48 | 0.11 | 0.3 | 2.41 | 0.11 | 3.37 | 0.3 | 0.19 | 0.59 |
| | 10 | 5.45 | 6.55 | 1.95 | 0.92 | 1.72 | 1.65 | 1.59 | 0.16 | 0.22 | 3.61 |
| | 11 | 7.15 | 9.94 | 3.96 | 1.72 | 2.57 | 3.37 | 2.61 | 0.31 | 1.11 | 6.92 |
| **7** | 0 | 3.2 | 1.26 | 0.55 | 2.41 | 3.47 | 4.21 | 8.43 | 4.38 | 4.32 | 4.04 |

| | | | | | | | | | | | |
|---|---|---|---|---|---|---|---|---|---|---|---|
| | 1 | 2.61 | 2.16 | 0.3 | 1.86 | 2.92 | 2.66 | 6.54 | 2.68 | 2.92 | 2.9 |
| | 2 | 3.26 | 4.06 | 0.29 | 1.14 | 2.36 | 2.29 | 4.9 | 1.36 | 1.65 | 3.05 |
| | 3 | 4.16 | 6.15 | 0.49 | 0.63 | 2.18 | 2.32 | 3.83 | 0.27 | 0.95 | 4.18 |
| | 8 | 7.63 | 1.68 | 0.32 | 4.24 | 5.57 | 0.56 | 6.99 | 4.01 | 3.82 | 2.77 |
| | 9 | 6.95 | 3.17 | 0.3 | 3.5 | 5.6 | 0.3 | 5.25 | 2.36 | 2.3 | 1.3 |
| | 10 | 6.63 | 4.86 | 0.99 | 3.09 | 5.23 | 0.23 | 3.54 | 1.01 | 1.13 | 1.16 |
| | 11 | 6.85 | 6.28 | 1.83 | 2.92 | 4.81 | 0.21 | 2.11 | 0.35 | 0.77 | 2.65 |
| **8** | 0 | 3.62 | 0.75 | 0.11 | 2.16 | 3.23 | 3.34 | 7.6 | 3.59 | 3.39 | 3.1 |
| | 1 | 2.62 | 3.23 | 0.11 | 1.25 | 2.16 | 1.31 | 4.73 | 1.19 | 1.51 | 1.94 |
| | 2 | 3.46 | 6.39 | 0.16 | 0.43 | 1.91 | 2.13 | 3.45 | 0.16 | 1.07 | 4.26 |
| | 3 | 5.46 | 9.68 | 1.97 | 1.98 | 3.12 | 4.08 | 4.21 | 0.2 | 3.62 | 7.51 |
| | 8 | 5.29 | 3.75 | 0.36 | 1.9 | 4.12 | 0.41 | 4.04 | 0.86 | 0.51 | 0.41 |
| | 9 | 5.84 | 6.33 | 2.18 | 1.58 | 2.7 | 0.6 | 0.83 | 0.33 | 0.31 | 2.94 |
| | 10 | 7.59 | 9.5 | 4.31 | 1.88 | 2.62 | 2.09 | 0.91 | 0.15 | 0.15 | 5.91 |
| | 11 | 9.8 | 12.9 | 7.12 | 2.9 | 3.78 | 4.56 | 1.65 | 1.25 | 0.26 | 9.37 |
| **9** | 0 | 2.44 | 4.14 | 0.32 | 3.18 | 3.89 | 3.76 | 6.94 | 3.31 | 3.93 | 4.52 |
| | 1 | 2.48 | 6.08 | 0.26 | 0.51 | 1.79 | 2.7 | 4.68 | 0.38 | 1.35 | 4.5 |
| | 2 | 4.68 | 9.41 | 1.51 | 0.25 | 1.36 | 4.39 | 4.26 | 0.11 | 2.29 | 6.9 |
| | 3 | 7.67 | 13.15 | 4.44 | 1.27 | 2.27 | 6.82 | 5.04 | 0.27 | 1.67 | 10.43 |
| | 8 | 3.46 | 4.41 | 0.1 | 1.13 | 2.46 | 2.29 | 4.77 | 1.1 | 1.51 | 3.3 |
| | 9 | 4.04 | 7.39 | 0.8 | 0.29 | 1.38 | 3.16 | 3.36 | 0.26 | 1.28 | 4.96 |
| | 10 | 5.63 | 10.57 | 2.81 | 0.74 | 1.7 | 4.12 | 3.46 | 0.22 | 1.87 | 7.83 |
| | 11 | 8.05 | 13.93 | 5.13 | 2.2 | 3.41 | 6.34 | 4.79 | 0.21 | 1.68 | 11.12 |
| **10** | 0 | 8.27 | 1.01 | 2.8 | 3.25 | 3.25 | 3.45 | 9.89 | 6.6 | 6.28 | 5.06 |
| | 1 | 5.69 | 0.35 | 0.36 | 0.23 | 0.34 | 0.81 | 6.62 | 3.18 | 2.87 | 1.66 |
| | 2 | 4.54 | 3.05 | 0.97 | 0.28 | 0.88 | 1.54 | 3.9 | 0.99 | 0.34 | 0.25 |
| | 3 | 5.91 | 6.53 | 3.09 | 0.81 | 1.51 | 2.42 | 2.41 | 1.53 | 0.16 | 3.07 |
| | 8 | 4.92 | 1.73 | 0.17 | 2.32 | 3.73 | 1.9 | 6.38 | 2.88 | 2.37 | 1.67 |
| | 9 | 4.56 | 4.15 | 0.2 | 0.84 | 2.9 | 0.2 | 2.95 | 0.1 | 0.1 | 0.9 |
| | 10 | 6.33 | 7.5 | 2.58 | 2.03 | 3.04 | 0.91 | 0.91 | 0.17 | 0.26 | 4.27 |
| | 11 | 8.58 | 11.09 | 5.14 | 3.62 | 4.57 | 2.62 | 1.45 | 0.25 | 1.55 | 7.86 |
| **11** | 0 | 5.41 | 0.83 | 1.85 | 0.53 | 0.18 | 2.34 | 7.17 | 4.03 | 3.57 | 2.52 |
| | 1 | 4.47 | 3.75 | 2.19 | 0.25 | 0.12 | 2.17 | 4 | 1.84 | 1.03 | 0.9 |
| | 2 | 6.33 | 7.22 | 4.03 | 0.21 | 0.22 | 2.7 | 2.67 | 1.29 | 0.21 | 3.85 |
| | 3 | 8.9 | 11.01 | 6.45 | 0.67 | 1.34 | 5.23 | 2.85 | 1.59 | 0.24 | 7.64 |

| | | | | | | | | | | | |
|---|---|---|---|---|---|---|---|---|---|---|---|
| | 8 | 5.81 | 3.06 | 1.16 | 1.65 | 3.54 | 0.87 | 4.16 | 0.8 | 0.34 | 0.32 |
| | 9 | 6.36 | 6.16 | 2.02 | 2.24 | 3.73 | 0.21 | 0.92 | 0.25 | 0.25 | 2.48 |
| | 10 | 7.94 | 9.41 | 3.72 | 4.3 | 5.18 | 0.17 | 0.17 | 0.17 | 1.64 | 5.95 |
| | 11 | 10.61 | 13.07 | 6.24 | 6.78 | 7.79 | 1.21 | 2.52 | 0.29 | 4.43 | 9.68 |
| **12** | 0 | 6.67 | 0.33 | 0.33 | 2.28 | 2.92 | 1.95 | 8.47 | 4.74 | 4.04 | 3.33 |
| | 1 | 4.9 | 1.95 | 0.36 | 0.9 | 2.37 | 1.02 | 4.92 | 1.35 | 0.76 | 0.23 |
| | 2 | 5.37 | 5.34 | 1.77 | 1.13 | 1.93 | 0.66 | 1.63 | 0.65 | 0.14 | 1.84 |
| | 3 | 7.79 | 9 | 4.34 | 2.19 | 3.08 | 2.23 | 1.53 | 0.17 | 0.41 | 5.49 |
| | 8 | 5.11 | 0.61 | 0.65 | 0.26 | 0.78 | 1.65 | 6.99 | 3.33 | 2.84 | 1.81 |
| | 9 | 4.3 | 3.67 | 1.28 | 0.13 | 0.57 | 1.99 | 3.67 | 0.88 | 0.19 | 0.49 |
| | 10 | 6.08 | 7.06 | 3.35 | 0.31 | 0.44 | 2.45 | 2.43 | 0.86 | 0.28 | 3.73 |
| | 11 | 8.05 | 10.67 | 5.55 | 0.99 | 1.8 | 4.86 | 2.75 | 0.84 | 0.28 | 7.36 |
| **13** | 0 | 2.69 | 1.86 | 0.38 | 2.39 | 3.33 | 4.06 | 8.12 | 4.16 | 4.25 | 4.06 |
| | 1 | 1.96 | 3.22 | 0.17 | 1.85 | 2.55 | 3.48 | 6.85 | 3.33 | 3.55 | 3.84 |
| | 2 | 2.07 | 5.01 | 0.23 | 1.27 | 1.97 | 3.62 | 6.04 | 2.09 | 2.37 | 4.52 |
| | 3 | 2.25 | 6.94 | 0.14 | 0.79 | 1.81 | 4.1 | 5.86 | 1.32 | 2.62 | 5.72 |
| | 8 | 3.53 | 3.59 | 0.39 | 1.67 | 2.92 | 1.89 | 4.98 | 1.67 | 2.15 | 2.64 |
| | 9 | 3.95 | 5.61 | 0.26 | 0.3 | 2.31 | 2.12 | 3.99 | 0.3 | 0.78 | 3.81 |
| | 10 | 4.49 | 7.73 | 1.33 | 0.87 | 2.06 | 3.6 | 4.02 | 0.29 | 1.83 | 5.52 |
| | 11 | 5.5 | 9.88 | 2.33 | 0.99 | 2.11 | 4.44 | 4.24 | 0.29 | 2.81 | 7.46 |
| **14** | 0 | 4.73 | 2.38 | 0.2 | 4 | 5.05 | 4.12 | 8.27 | 4.61 | 4.55 | 4.09 |
| | 1 | 3.75 | 3.69 | 0.22 | 2.5 | 3.66 | 1.28 | 4.96 | 1.28 | 2.23 | 2.11 |
| | 2 | 4.98 | 6.75 | 0.67 | 1.77 | 3.43 | 1.28 | 2.73 | 0.26 | 1.83 | 3.92 |
| | 3 | 7.4 | 10.24 | 3.56 | 3.79 | 4.84 | 2.11 | 2.32 | 0.26 | 3.02 | 7.33 |
| | 8 | 6.05 | 3.61 | 0.16 | 2.84 | 5.03 | 0.54 | 5 | 1.86 | 1.64 | 0.86 |
| | 9 | 6.42 | 6.03 | 1.69 | 2.38 | 4.19 | 0.2 | 1.8 | 0.2 | 0.2 | 2.51 |
| | 10 | 7.97 | 9.28 | 4.01 | 3.94 | 4.74 | 0.17 | 0.19 | 0.17 | 1.17 | 5.85 |
| | 11 | 10.53 | 12.88 | 6.83 | 5.69 | 6.64 | 1.87 | 1.08 | 0.2 | 2.5 | 9.42 |
| **15** | 0 | 3.46 | 4.91 | 2.9 | 1.74 | 0.19 | 4.93 | 5.99 | 3.23 | 2.89 | 3.59 |
| | 1 | 2.07 | 4.33 | 0.49 | 0.13 | 0.15 | 3.23 | 5.13 | 1.5 | 1.61 | 3.06 |
| | 2 | 1.88 | 4.91 | 0.24 | 1.25 | 2.18 | 2.76 | 5.24 | 1.08 | 1.9 | 4.17 |
| | 3 | 3.56 | 6.88 | 0.17 | 3.8 | 4.8 | 3.52 | 6.78 | 2.38 | 4.3 | 6.08 |
| | 8 | 6.58 | 4.58 | 0.26 | 3.79 | 5.86 | 0.71 | 5.08 | 1.83 | 2.44 | 2.09 |
| | 9 | 6.32 | 5.45 | 0.99 | 2.7 | 4.79 | 0.15 | 3.04 | 0.47 | 1.24 | 2.01 |

| | | | | | | | | | | | |
|---|---|---|---|---|---|---|---|---|---|---|---|
| | 10 | 7.23 | 7.21 | 2.53 | 3.34 | 4.67 | 0.24 | 1.48 | 0.06 | 0.86 | 3.73 |
| | 11 | 8.48 | 9.47 | 4.24 | 4.68 | 5.49 | 0.04 | 0.49 | 0.04 | 1.38 | 5.93 |
| **16** | 0 | 5.38 | 1.53 | 0.24 | 3.12 | 4.42 | 2.97 | 7.64 | 4.13 | 3.68 | 2.97 |
| | 1 | 4.52 | 2.36 | 0.13 | 1.95 | 3.7 | 0.69 | 5.29 | 1.75 | 1.4 | 0.67 |
| | 2 | 5.05 | 4.28 | 0.7 | 1.23 | 3.1 | 0.23 | 2.96 | 0.15 | 0.14 | 0.63 |
| | 3 | 6.94 | 6.7 | 2.97 | 2.77 | 3.66 | 0.59 | 0.71 | 0.95 | 0.29 | 3.09 |
| | 8 | 7.17 | 3.25 | 1.23 | 3.18 | 5.47 | 0.28 | 4.35 | 1.75 | 1.69 | 0.99 |
| | 9 | 7.06 | 5.6 | 2.06 | 3.11 | 4.94 | 0.24 | 2.25 | 0.99 | 1.04 | 1.95 |
| | 10 | 8.26 | 7.96 | 3.56 | 4.19 | 5.33 | 0.24 | 0.87 | 0.57 | 0.73 | 4.2 |
| | 11 | 9.66 | 10.43 | 5.27 | 5.89 | 6.68 | 0.25 | 0.51 | 0.79 | 2.4 | 6.73 |

Contacts 0-3 represent the left hemisphere lead, and 8-11 the right side with the lower contacts being the most proximal ones. AD – anterodorsal thalamic nucleus, AM – anteromedial thalamic nucleus, AV – anteroventral thalamic nucleus, CL – central lateral thalamic nucleus, MDpc – medial dorsal thalamic nucleus pars parvocelullaris, MTT – mammillothalamic tract, VApc - ventral anterior thalamic nucleus pars parvocellularis, VLa – ventral lateral thalamic nucleus pars anterior, VLpd – ventral lateral thalamic nucleus posterior dorsal, VLpv - ventral lateral thalamic nucleus posterior ventral.

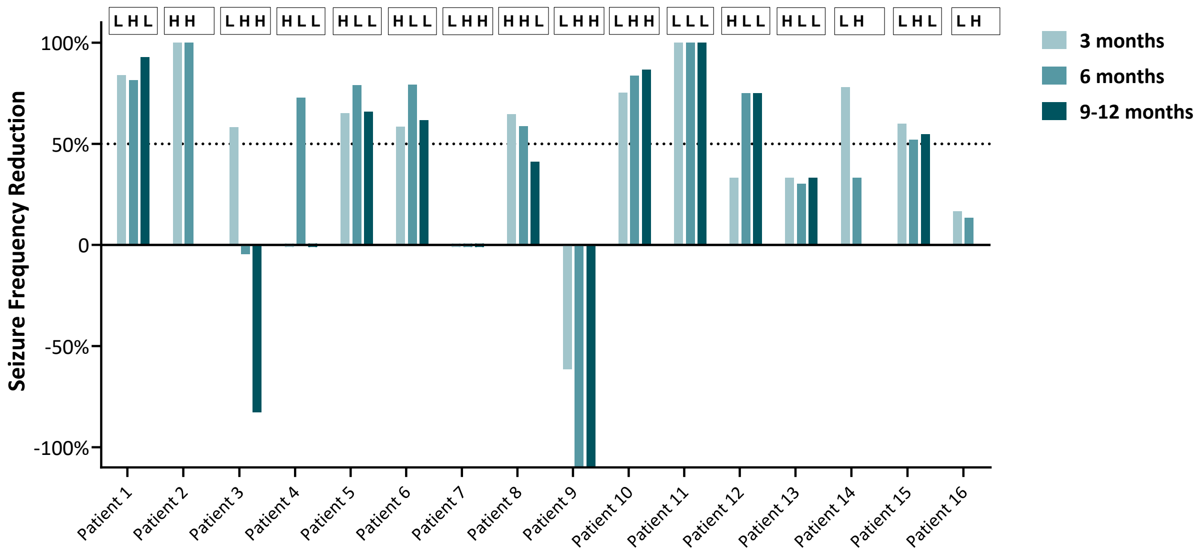

Supplementary Figure 1 – **Individual seizure frequency reduction at each assessment.** Above each column is the stimulation parameter that the patient was on for that assessment. H = iHFS parameters that consist of 145 Hz frequency, 90 µsec pulse width, and cycling 1 min on and 5 min off: L = cLFS parameters consisting of 7 Hz frequency, 200 µsec pulse width, and continuous stimulation.

**Assessment of priming effect**

Priming effect refers to the facilitation of network modulation (either enhancing or blocking the excitability of a system) after an initial stimulus.[3] This concept mainly applies to motor network physiology. However, priming in epilepsy, which is a strictly theoretical concept, could be seen as an initial set of parameters that facilitates epileptic network inhibition for the subsequent set; i.e. the second set seems more effective than the first at seizure reduction, although some of its efficacy is in part due to the initial set of parameters paving the way.

To assess priming we compared the median SFR at 6 months (second assessment) in patients that started on iHFS first and the switch to cLFS vs those who did the opposite. Median SFR was not different in the seven individuals that were started on iHFS (72%, IQR 30 – 79) vs the seven individuals that were started on cLFS (33%, IQR -1 to 84) at the time of second assessment (p = .7) (Supplementary Figure 2).

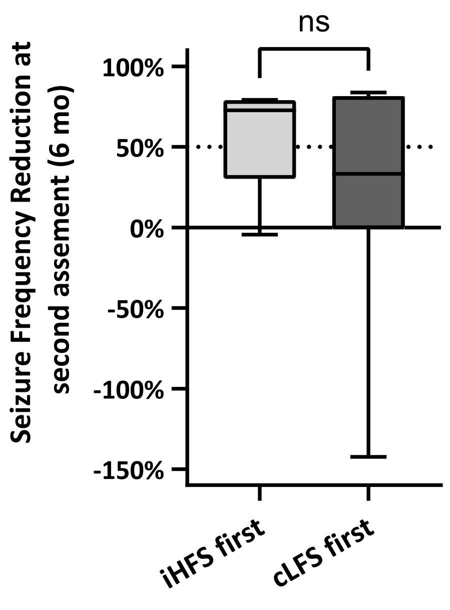

Supplementary Figure 2 – **Comparison of seizure frequency reduction at 6 months (second assessment) in patients who started with iHFS parameters first vs. patients**

**who started with cLFS parameters first.** cLFS, continuous low frequency stimulation, iHFS, intermittent high frequency stimulation; ns = not statistically significant

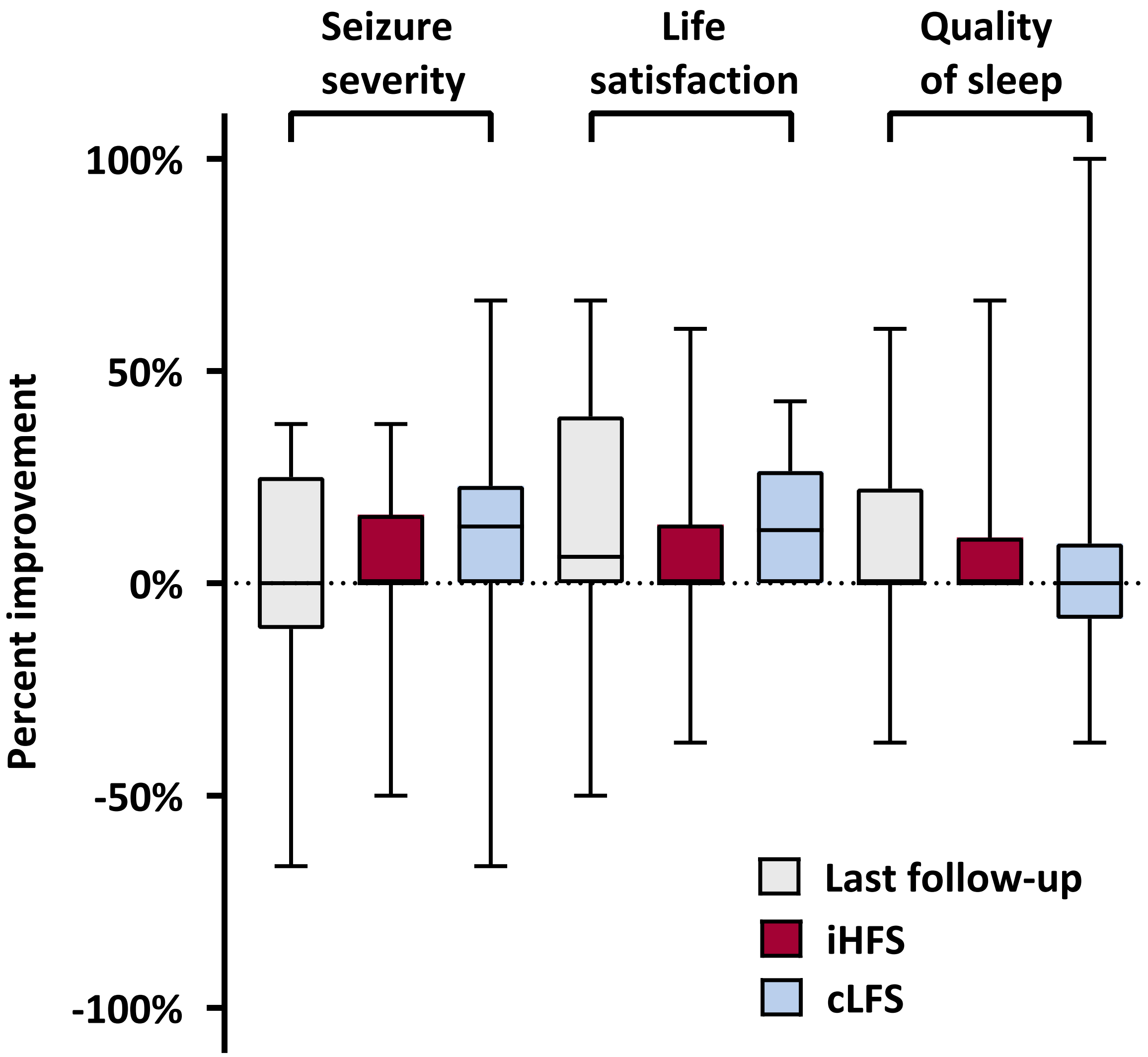

Supplementary Figure 3 - **Improvement of patient perceived outcomes**. Using 1-10 analog numeric scales for their assessment, there was no significant improvement in seizure frequency, life satisfaction or quality of sleep. Abbreviations: cLFS, continuous low frequency stimulation, iHFS, intermittent high frequency stimulation.

**Implantation related adverse events and stimulation related side effects assessment**

To assess DBS hardware implantation-related adverse events (AE) (Supplementary table 1) and stimulation-related side effects (SE) (Supplementary Table 2), patients were instructed to fill out the following forms: Form 1 at the randomization visit and Form 2 was filled out at every visit for seizure frequency evaluation. When forms were not filled out by the participants, the clinician documented any suspicious AE/SE in the visit note. Supplementary Figure 4.

Severity definitions:

- Mild AE/SE were defined as those that required no specific medical intervention such SE that resolved spontaneously with no intervention or with over-the-counter medications such as acetaminophen use for pain.
- Moderate AE/SE were defined as those requiring minimal intervention such as stimulation parameter modification, evaluation by clinician (telemedicine or in-person), local intervention, or specific monitoring.
- Severe AE/SE were defined as those requiring hospitalization and/or surgical intervention.

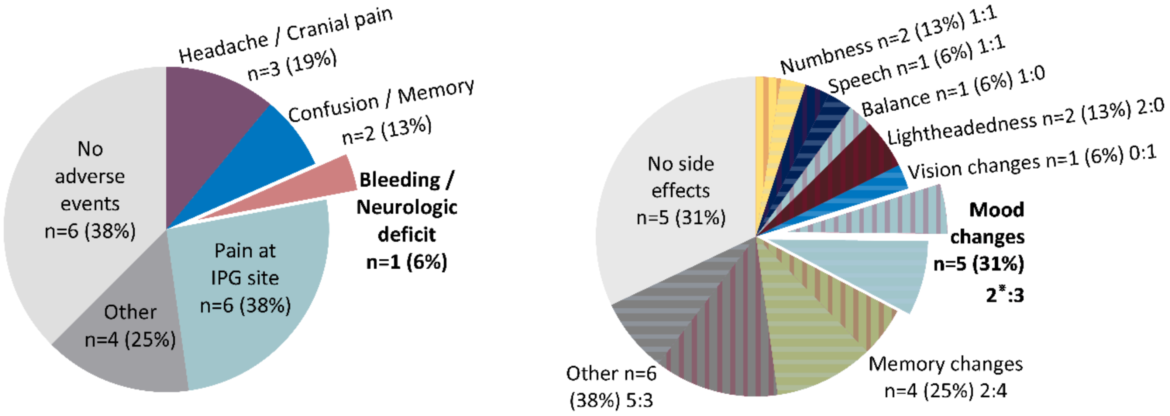

Supplementary Figure 4 - **Implant related adverse events (AEs), left panel, and stimulation related side effects, (SEs), right panel**. The pies represent the proportion of patients that developed the AE/SE as patients could have more than one AE/SE simultaneously. The percentages demonstrate how common the AE/SE was for any given patient. AE/SE that were classified as moderate as they required dedicated monitoring or intervention are in bold. For the right graph, the horizontal blue stripped pattern represents cLFS-related SEs, while the red vertical stripped pattern represents iHFS-related SEs. Mood side effects were present in both iHFS (n=2) and cLFS (n=3), however only the iHFS-related cases required an intervention, while the three cLFS-related cases resolved spontaneously.

Supplementary table 2 - Hardware implantation-related adverse events

| Device implantation related AE | AE severity | Description |
|---|---|---|
| Headache / Cranial pain | Mild | • Patient 13 had mild headaches over the two weeks following implant that resolved with acetaminophen<br>• Patient 15 had pain around the lead entry sites when raising eyebrows or squinting that required no intervention or evaluation and subsided spontaneously<br>• Patient 14 referred constant headache at lead entry sides, especially when sleeping on their back |
| Confusion/Memory | Mild | • Patient 1 had intermittent word finding difficulties in the first few days after implant that resolved by the time of randomization<br>• Patient 14 had mild difficultly recalling recent memories, information, and finding words that resolved by day two post-surgery |
| Bleeding | Moderate | • Patient 6 had blood products around the left lead and mild, transient right facial droop and right upper extremity drift that resolved in 24 hours and delayed discharge for one day. |
| Pain at IPG implant site or wire length (neck) | Mild | • Six patients (1,4,7,11,13,14) had mild tenderness and swelling around the IPG incision that either self-resolved spontaneously or with acetaminophen.<br>• Patient 13 noted stiffness around the neck wiring that caused mild pain overnight after surgery |
| Other | Mild | • Patient 8 noted dizzy spells after sharply turning their head since DBS, irrespective of stimulation<br>• Patient 14 noted numbness in the top of their scalp that self-resolved<br>• Patient 15 noted exacerbation of phonophobia and headaches triggered by high-pitched sounds that resolved with acetaminophen. |
| | Moderate | • Patient 2 had a cluster of four convulsive seizures right in the immediate postoperative period that delayed discharge for 24 hours and prompted the initiation of lacosamide |

Abbreviations: AE, adverse events; DBS, deep brain stimulation; IPG, implantable pulse generator.

Supplementary Table 3 - Stimulation-related side effects

| | Severity and description of SE while on iHFS | Severity and description of SE while on cLFS |
|---|---|---|
| Numbness | • **Mild -** Patient 12 had tingling/vibration around the area of the wire on the right side of the back of their neck once while singing, and a slight burning sensation on their right chest over IPG that follows the wire up to the right collarbone weekly during the day. The burning sensation did not hurt, but it was noticeable. | • **Mild -** Patient 3 had a transient uncomfortable numb sensation during programing that subsided later in the day. |
| Speech | • **Mild -** Patient 16 noticed intermittent worsening of their baseline word finding difficulties. | • **Mild -** Patient 16 noticed intermittent worsening of their baseline word finding difficulties. |
| Balance | • **Mild -** Patient 16 noticed mild worsening of their baseline balance issues. | - |
| Lightheadedness | • **Mild -** Patient 7 had a single instance of lightheadedness after switching to iHFS. This resolved spontaneously the same day.<br>• **Mild -** Patient 16 reported vague dizziness. | - |
| Vision changes | - | • **Mild -** Patient 8 noted trouble focusing on objects early in the morning and late at night. |
| Mood changes | • **Moderate -** Patient 3 developed depressive mood with suicidal ideation after increasing current to 2.0 mA per cathode. This resolved after decreasing current to 1.7 mA.<br>• **Moderate -** Patient 12 had multiple instances of marked déjà vu and panic attacks after increasing current to 2 mA, which prompted turning off the device for two days. cLFS was later started at 0.5 mA per cathode, and they did not experience the SE again. | • **Mild -** Patient 6 became angrier towards family, although this was seen as a favorable sign as the patient was more interactive after having less seizures.<br>• **Mild -** Patient 16 reported increased anger and low mood related to IPG site discomfort.<br>• **Mild -** Patient 11 noticed mild depressive symptoms since starting stimulation. This was attributed to a social stressor (college course) as the symptoms resolved after they dropped out from the course. |
| Memory changes | • **Mild -** Patient 12 had mild worsening of working memory that required the use of calendar and | • **Mild -** Patient 1 noted mild short term memory issues compared to before. |

| | | |
|---|---|---|
| | reminders when initiating stimulation with iHFS.<br><br>• **Mild -** Patient 16 noticed worsening of their baseline limited short term recall. | • **Mild -** Patient 8 noted trouble remembering conversations that was overcome with prompting.<br><br>• **Mild -** Patient 12 reported further memory decline after switching from iHFS to cLFS that was overcome by using reminder apps.<br><br>• **Mild -** Patient 16 noticed worsening of their baseline limited short term recall that was not different from the one reported while on iHFS. |
| Other | • **Mild -** Patient 1 had transient shooting pain around the IPG.<br><br>• **Mild -** Patient 7 noted headaches while on iHFS.<br><br>• **Mild -** Patient 13 felt "jumpy" for about 10 min after initiating stimulation with iHFS.<br><br>• **Mild -** Patient 14 noticed intermittent nausea while on iHFS irrespective of current changes.<br><br>• **Mild** – Patient 16 reported occasional pain in the IPG pocket at times extending to the neck | • **Mild -** Patient 13 felt "jumpy" for about 10 min after changing from iHFS to cLFS.<br><br>• **Mild -** Patient 14 noticed transient nausea only with current changes.<br><br>• **Mild -** Patient 15 developed a prodrome before their seizures described as an "electric wave" which they had not experienced prior to DBS. |

Abbreviations: DBS, deep brain stimulation; iHFS, high frequency stimulation; IPG, implantable pulse generator; LCM, lacosamide; cLFS low frequency stimulation; mA, milliamperes; min, minutes; SE, side effect

Form 1 - Pre-Stimulation Questionnaire

| **Stimulation Activation Visit** | |
|---|---|
| **Have you experienced any of the following since implantation? If you have, please rate the severity, and provide a brief description and an estimated date of onset:** | |
| **1. Infection**<br>Date of Onset:<br>Brief Description: | ☐ 1 – Mild<br>☐ 2 – Moderate<br>☐ 3 – Severe<br>☐ 4 – N/A |
| **2. Headache**<br>Date of Onset:<br>Brief Description: | ☐ 1 – Mild<br>☐ 2 – Moderate<br>☐ 3 – Severe<br>☐ 4 – N/A |
| **3. Confusion**<br>Date of Onset:<br>Brief Description: | ☐ 1 – Mild<br>☐ 2 – Moderate<br>☐ 3 – Severe<br>☐ 4 – N/A |
| **4. Difficulty concentrating**<br>Date of Onset:<br>Brief Description: | ☐ 1 – Mild<br>☐ 2 – Moderate<br>☐ 3 – Severe<br>☐ 4 – N/A |
| **5. Device deficiencies**<br>Date of Onset:<br>Brief Description: | ☐ 1 – Mild<br>☐ 2 – Moderate<br>☐ 3 – Severe<br>☐ 4 – N/A |
| **6. Temporary pain, swelling and the implant site**<br>Date of Onset:<br>Brief Description: | ☐ 1 – Mild<br>☐ 2 – Moderate<br>☐ 3 – Severe<br>☐ 4 – N/A |
| **7. Other, please describe:**<br>Date of Onset:<br>Brief Description: | ☐ 1 – Mild<br>☐ 2 – Moderate<br>☐ 3 – Severe<br>☐ 4 – N/A |

Form 2 - Post-Stimulation Activation Questionnaire

| **Follow Up Visits 1, 2, and 3** | |
|---|---|
| **Have you experienced any of the following since the stimulation initiation (month 0) visit? If you have, please rate the severity, and provide a brief description and an estimated date of onset:** | |
| **1. Numbness or tingling sensations**<br>Date of Onset:<br>Brief Description: | ☐ 1 – Mild<br>☐ 2 – Moderate<br>☐ 3 – Severe<br>☐ 4 – N/A |
| **2. Muscle tightness of the face or arm**<br>Date of Onset:<br>Brief Description: | ☐ 1 – Mild<br>☐ 2 – Moderate<br>☐ 3 – Severe<br>☐ 4 – N/A |
| **3. Speech problems**<br>Date of Onset:<br>Brief Description: | ☐ 1 – Mild<br>☐ 2 – Moderate<br>☐ 3 – Severe<br>☐ 4 – N/A |
